# Persistence of Neuropsychiatric Symptoms and Dementia Prognostication: A Comparison of Three Operational Definitions of Mild Behavioral Impairment

**DOI:** 10.1101/2023.04.08.23287454

**Authors:** Dylan X. Guan, Eric E. Smith, G. Bruce Pike, Zahinoor Ismail

## Abstract

**INTRODUCTION:** This study compares three operational definitions of mild behavioral impairment (MBI) in the context of MBI prevalence estimates and dementia risk modeling.

**METHODS:** Participants were dementia-free older adults (n=13701) from the National Alzheimer’s Coordinating Center. Operational definitions of MBI were generated based on neuropsychiatric symptoms at one (OV), two-consecutive (TCV), or >2/3 (TTV) of dementia-free study visits. Definitions were compared in prevalence and in Cox regressions using MBI to predict incident dementia.

**RESULTS:** OV MBI was the most prevalent (54.4%), followed by TCV (32.3%) and TTV (26.7%) MBI. However, OV MBI had the lowest rate of incident dementia (HR=2.54, 95%CI: 2.33–2.78) and generated poorer model metrics than TCV MBI (HR=4.06, 95%CI: 3.74–4.40) and TTV MBI (HR=5.77, 95%CI: 5.32–6.26).

**DISCUSSION:** Case ascertainment with longer timeframe MBI operational definitions may more accurately define groups at risk of dementia in datasets lacking tools designed to detect MBI.

**HIGHLIGHTS:**

- Mild behavioral impairment (MBI) can identify older adults at risk of dementia.
- Neuropsychiatric symptom (NPS) assessment tools can be proxy measures for MBI.
- Hazard for dementia was highest for MBI defined by NPS presence at >2/3 of visits.

## 1. BACKGROUND

Mild behavioral impairment (MBI) is a syndrome that identifies older adults at high risk for neurodegenerative disease using neuropsychiatric symptoms (NPS).^1^ These NPS include decreased motivation (apathy), affective dysregulation (mood and anxiety), impulse dyscontrol (agitation and aggression), social inappropriateness, and abnormal perception or thought content (delusions and hallucinations; psychosis). NPS qualify as symptoms of MBI when they (1) emerge *de novo* in later life and persist at least intermittently for ≥6 months; (2) cannot be explained by previous or current psychiatric conditions; and (3) occur in older adults without dementia. These criteria improve specificity for the NPS identified by MBI representing early-stage neurodegenerative disease as a complementary behavioral analog to late-life emergent cognitive symptoms.^2^ As such, MBI has the potential to improve early detection of dementia risk, thereby facilitating research into early neurodegenerative disease mechanisms and the development of preventative or disease-modifying treatments.^3,4^

The Mild Behavioral Impairment Checklist (MBI-C) was developed specifically to capture NPS that meet the criteria for MBI.^5^ Studies that used the MBI-C demonstrated poorer cognition and greater β-amyloid (Aβ) and tau pathology, medial temporal lobe atrophy, and Alzheimer’s disease (AD) genetic risk in older adults with MBI.^6-13^ However, given the relative novelty of the Alzheimer’s Association International Society to Advance Alzheimer’s Research and Treatment (ISTAART-AA) MBI criteria and the MBI-C, which were published in 2016 and 2017, respectively, datasets with abundant MBI-C data are not yet widely available. To address this issue, other existing measures of NPS, most commonly the Neuropsychiatric Inventory (NPI) and its derivatives, have been used as a proxy measure of MBI with algorithms and specific data cleaning procedures.^14,15^ This approach has generated novel insight into MBI and its relationship to dementia features that would have been overlooked until more frequent MBI-C endorsement. Longitudinally, MBI, defined using the NPI or NPI Questionnaire (NPI-Q), has consistently been associated with a greater risk for cognitive decline and incident dementia,^15-18^ and a lower rate of reversion from mild cognitive impairment (MCI) to normal cognition in a sample of participants from the National Alzheimer’s Coordinating Center (NACC).^19^ In other datasets such as the Alzheimer’s Disease Neuroimaging Initiative and MEMENTO, where only NPI or NPI-Q data are available, MBI was linked to more severe neurodegenerative disease pathology, including lower plasma Aβ42/Aβ40 and higher phosphorylated tau and neurofilament light, in addition to greater white matter hyperintensity volume and entorhinal cortex atrophy.^20-23^ Finally, in the Canadian Comprehensive Assessment of Neurodegeneration and Dementia study, MBI derived from the NPI-Q was associated with worse gait, hearing loss, and frailty, all of which are non-cognitive markers of dementia.^24-27^

The use of other NPS assessment tools as a proxy measure for MBI will likely continue to produce important findings related to MBI, especially in datasets with longitudinal data that preceded the publication of the MBI-C. Yet, current methods used to operationally define MBI using proxy tools face limitations, potentially hindering accurate differentiation between MBI and non-MBI NPS. In older adults with MCI, only MBI was associated with a lower likelihood of reversion to normal cognition, whereas non-MBI NPS showed no difference compared to no NPS.^19^ This finding suggests that distinguishing MBI from non-MBI NPS is essential to fully understand and make use of MBI as a tool for neurodegenerative disease research and for sample enrichment.^4^ One issue with using the NPI and its derivatives for MBI is the reference timeframe by which the instruments assess NPS, which spans one month.^28-30^ In contrast, the MBI-C reference frame is six months, consistent with the symptom persistence criterion for MBI.^5^ Consequently, studies that operationally define MBI using NPI or NPI-Q data from a single visit risk misclassifying as MBI, NPS that are transient and potentially arising due to non-neurodegenerative etiologies. Furthermore, estimates of MBI prevalence in samples of older adults without dementia are considerably higher when using the NPI or NPI-Q (28-85%)^14,26,31^ compared to the MBI-C (6-14%)^6,7,32,33^ at a single visit, suggesting low specificity when operationally defining MBI this way. More recent studies have operationally defined MBI based on the presence of NPS across two consecutive visits, often separated by approximately six or twelve months, to meet the persistence criterion for MBI.^17,19,20,22^ However, due to the 1-month timeframes of the NPI and NPI-Q instruments, this method still risks misclassifying participants with transient NPS as having MBI. A new operational definition to identify older adults with MBI more accurately, and to distinguish MBI from non-MBI NPS, is warranted.

This study proposes a new method to operationalize MBI in datasets with retrospective longitudinal data that have not implemented the MBI-C. To demonstrate its utility in real datasets, we compare the novel method to previously used operational definitions of MBI in terms of MBI prevalence estimates and dementia modeling performance in NACC data.

## 2. METHODS

### 2.1. Study Design

Data were obtained from the NACC Uniform Dataset (NACC-UDS). Forty-five Alzheimer’s Disease Research Centers (ADRCs) funded by the National Institute on Aging contributed to this dataset between 2005-2022. Each ADRC recruited and collected data, including for NPS, on participants with or without dementia approximately annually. All ADRCs obtained informed consent from their participants and received ethics approval from their institutions prior to submitting data to NACC. More detailed information about the NACC-UDS and its data collection procedures has been published elsewhere.^34-37^

### 2.2. Participants

Longitudinal data for 45100 participants were available in the NACC-UDS. Participants had normal cognition (CN), subjective cognitive decline (SCD), MCI, or dementia. As shown in Figure 1, participants were excluded if they: (1) were <50 years old, consistent with MBI criteria,^1^ n=1069; (2) reported a history of relevant psychiatric conditions, n=14556; (3) failed to report years of education completed, n=244; or (4) were missing NPI-Q data, n=910. Applying the exclusion criteria resulted in 28321 participants who were eligible for analysis. Relevant psychiatric conditions included posttraumatic stress disorder, bipolar disorder, schizophrenia, obsessive-compulsive disorder, remote anxiety or depression, which preclude MBI diagnosis.^1^ Excluded neurological conditions included Down syndrome, and Huntington disease.

**Figure 1.**
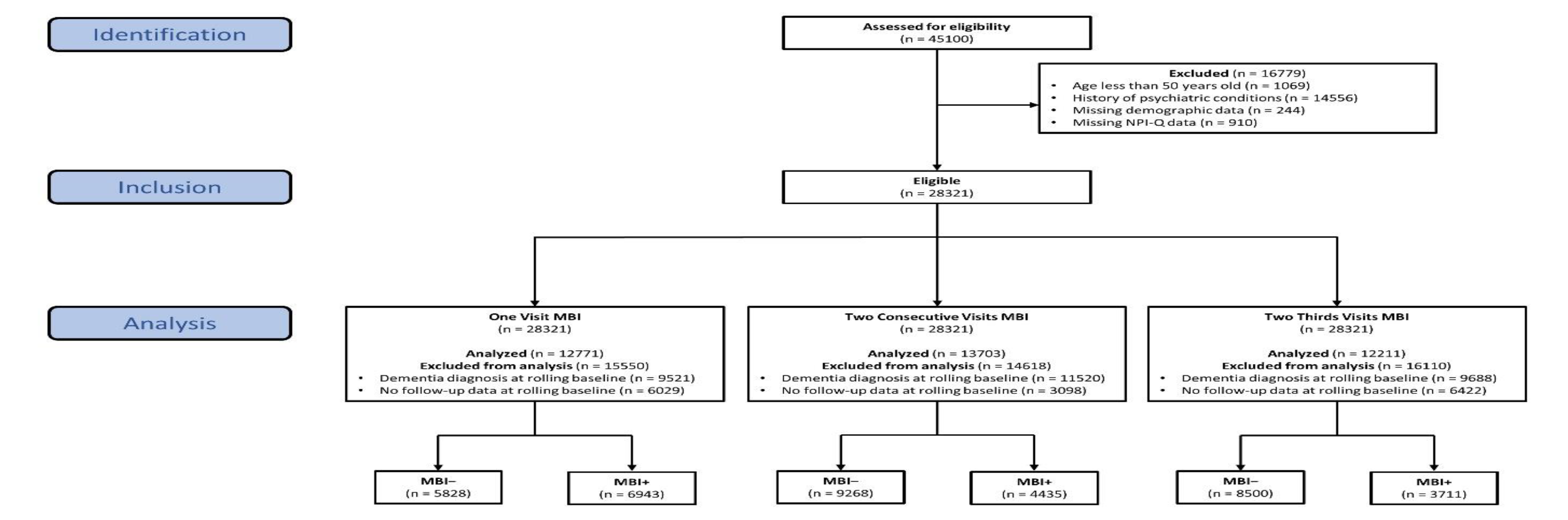
Participant Flow Diagram. Data came from the National Alzheimer’s Coordinating Center (NACC). Relevant psychiatric conditions used for exclusion included posttraumatic stress disorder, bipolar disorder, schizophrenia, obsessive-compulsive disorder, remote anxiety or depression, Down syndrome, and Huntington disease. Abbreviations: NPI-Q, Neuropsychiatric Inventory Questionnaire; MBI, mild behavioral impairment. **\***Color should be used for this figure

### 2.3. MBI Operational Definitions

The presence and severity of NPS were measured using the NPI-Q.^28,30^ Briefly, informants rated the presence and severity of NPS on a scale from 1-3 over the previous month. A published algorithm was used to derive MBI symptom severity score from 10/12 NPI-Q items (excluding sleep and appetite) based on the ISTAART-AA diagnostic criteria for MBI.^14^ Specifically, the decreased motivation domain was derived from the NPI-Q apathy item; emotional dysregulation from the NPI-Q depression, anxiety, and elation items; impulse dyscontrol from the NPI-Q agitation, irritability, and aberrant motor behavior items; social inappropriateness from the NPI-Q disinhibition item; and psychosis from the NPI-Q delusions and hallucinations items. The total MBI symptom severity (range=0–30) was computed as the sum of all five MBI domain scores, with higher scores indicating more severe MBI symptoms.

Three operational definitions of MBI were explored. The first, termed One Visit (OV) MBI, classified participants as having MBI if they had a total MBI symptom severity score ≥1 at a single dementia-free study visit. The second operational definition, termed Two Consecutive Visits (TCV) MBI, classified participants as having MBI if they had a total MBI symptom severity score ≥1 for two consecutive dementia-free study visits. The third operational definition, termed Two-Thirds Visits (TTV) MBI, classified participants as having MBI if the participant had a total MBI symptom severity score ≥1 at a dementia-free study visit and for >2/3 of all subsequent dementia-free study visits, when including the initial visit with NPS. To account for participants who developed MBI over the course of their observational period in NACC, participants were categorized MBI+ if they developed MBI at any visit and MBI– if they never developed MBI at any visit. A rolling baseline was applied to MBI+ participants; their baseline was shifted to the first visit they were classified as MBI+ and all prior visits were considered pre-MBI. Correspondingly, time to dementia was calculated as the difference in years between the first dementia visit and the rolling baseline visit. As the baseline was shifted for MBI+ participants, additional exclusion criteria were applied to each set of analyses involving one of the three MBI operational definitions: participants were excluded for having a clinical dementia diagnosis (OV n=9521, TCV n=11520, TTV n=9586) or no follow-up data (OV n=6029, TCV n=3098; TTV n=5034) at the rolled baseline. These steps resulted in a final sample of 12771 participants for the OV analysis, 13703 for the TCV analysis, and 13701 for the TTV analysis. Example scenarios illustrating differences between the three MBI operational definitions in the MBI classification of participants and how dementia-free survival time was calculated using the rolling baseline, if appropriate, are shown in Figure 2. Simulated data accompanied by a data dictionary and the R code used to generate each MBI operational definition in this study are available at https://osf.io/k75rg for those interested in exploring MBI case ascertainment scenarios or implementing these operational definitions in their own studies.

**Figure 2.**
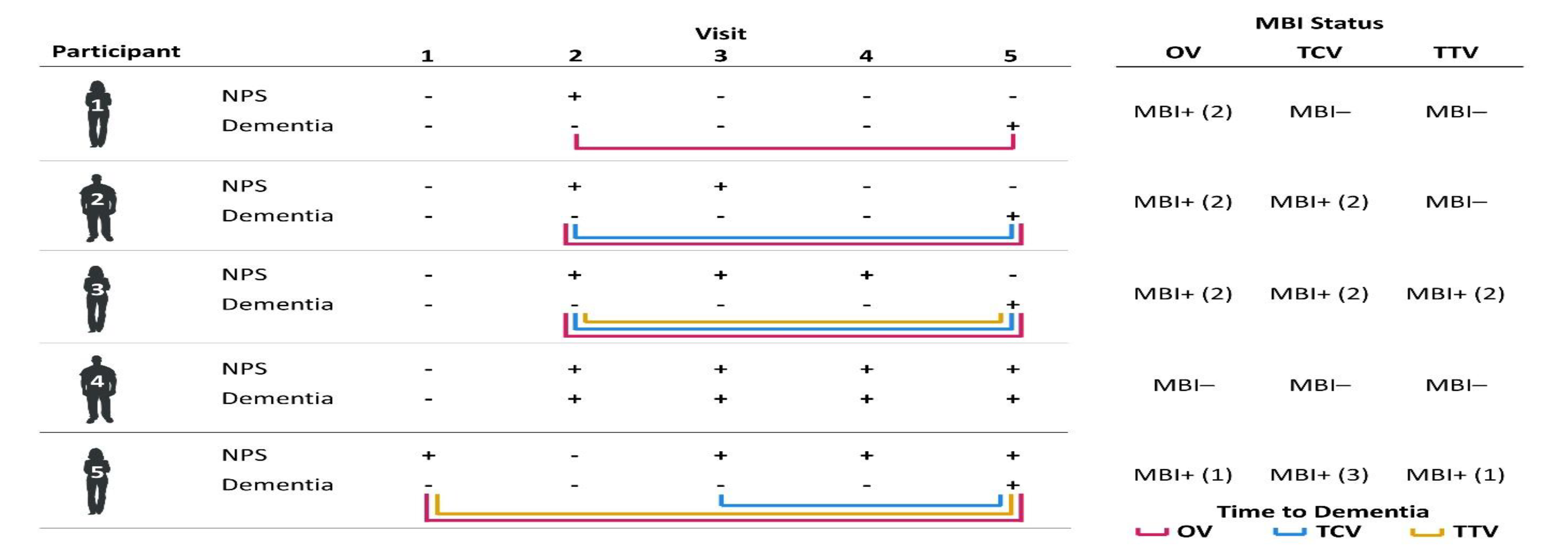
Example Case Ascertainment for Three MBI Operational Definitions. Positive/negative signs indicate the presence/absence, respectively, of NPS or a dementia diagnosis at a given study visit for a participant. Numbers in parentheses in the right table indicate the study visit at which a participant was first defined as having MBI, if applicable. Colored lines indicate the time between MBI classification, if applicable, and the first dementia visit. Abbreviations: NPS, Neuropsychiatric symptoms; MBI, mild behavioral impairment; OV, One Visit MBI operational definition; TCV, Two Consecutive Visits MBI operational definition; Two-Thirds Visits MBI operational definition. **\***Color should be used for this figure

### 2.4. Statistical Analysis

Descriptive statistics including means, standard deviations (SDs), ranges, medians, interquartile ranges (IQRs), and percentages were used to describe participant demographic and clinical characteristics. Between-group comparisons for MBI– and MBI+ participants, defined using each of the three operational definitions, were conducted using independent samples t-tests for continuous variables or chi-square tests for categorical variables. To determine potential trends in missing NPI-Q data, demographic comparisons were also conducted between participants eligible for analysis and those excluded for missing NPI-Q data. Prevalence estimates of MBI were determined by dividing the number of MBI+ participants by the total sample size, and then multiplying by 100 to obtain a percentage.

Dementia-free survival was compared between MBI groups using Kaplan-Meier survival estimates. To extract measures of effect size (hazard ratio; HR), Cox proportional hazards regression models were conducted with MBI status as the predictor variable and incident dementia as the outcome variable. These Cox regressions were first run as univariable models to compare the effect size corresponding to each MBI operational definition independently of covariates. Subsequently, multivariable Cox regression models adjusted for age, sex, education, and cognitive status (CN/SCD or MCI) to control for other factors that may contribute to the rate of incident dementia. Schoenfeld and Martingale residuals were evaluated to verify that the proportional hazards and linearity assumptions, respectively, were met for predictors in each Cox regression model, as appropriate.

To compare the three operational definitions of MBI for dementia modeling performance, we extracted the concordance index (C-index) and Akaike’s information criterion (AIC) for each model. The C-index is a commonly used evaluation metric for the predictive ability of survival models with higher values indicating greater prediction accuracy,^38^ and the AIC may be used to compare model performance between non-nested models in the same data with lower values indicating better model fit.^39^ Sensitivity analyses were used to compare MBI operational definitions using higher MBI total score cut-offs of ≥2 and ≥3. All analyses employed a statistical significance threshold of p < .05 and were conducted on R version 4.0.2.^40^ Data access was provided by NACC after submitting an approved research proposal.

## 3. RESULTS

Baseline participant characteristics for each sample are summarized in Table 1. Across all three cohorts, participants were approximately 57% female, 73 years old (SD=9, range=50–104), and had completed 16 years of education (SD=3, range=0–30). The most common cognitive statuses were CN (63%), followed by MCI (31%) and SCD (6%). The median follow-up time was approximately 3 years (IQR=2–6) and 18% of participants progressed to dementia during follow-up. Comparisons between MBI– and MBI+ participants, according to each MBI operational definition, can be found in Tables S1-S3. Participants that were missing NPI-Q data were more likely to be older and less likely to have cognitive impairment, than participants with complete NPI-Q data [Table S4].

**Table 1.**
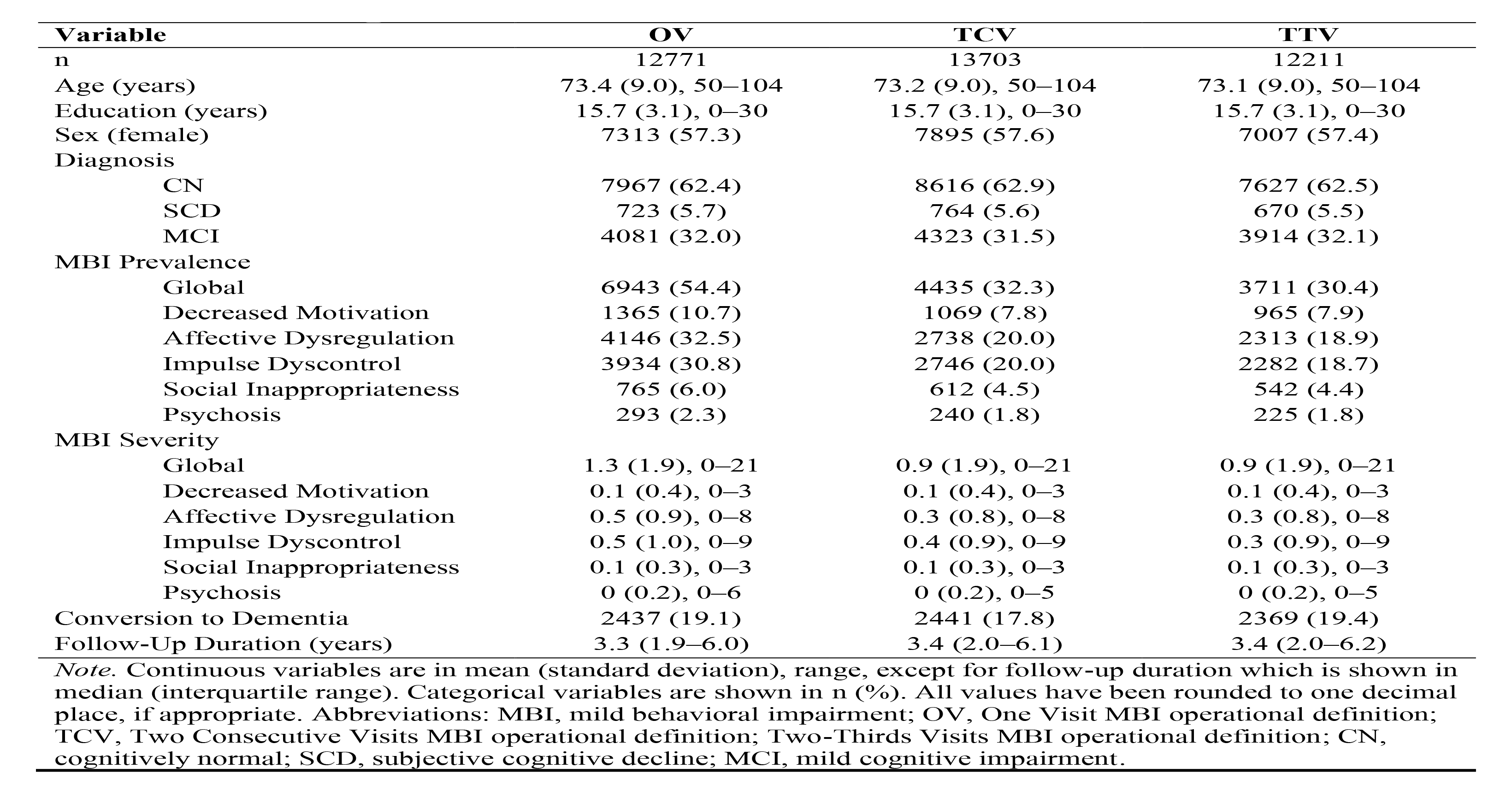
Baseline Participant Characteristics

The prevalence of MBI was highest using the OV operational definition (54.4%), followed by TCV (32.3%), and lowest using TTV (30.4%). Regardless of operational definition, affective dysregulation and impulse dyscontrol tended to be the most prevalent MBI domains followed in order by decreased motivation, social inappropriateness, and psychosis. Kaplan-Meier survival estimates revealed that 25.9% of older adults with OV MBI progressed with an estimated median time to dementia of 11.77 years (95%CI: 11.01–13.02), as shown in Figure 3. In contrast, 33.0% of older adults with TCV MBI progressed with a median time to dementia of 7.28 years (95%CI: 6.78–8.51), and 39.8% of older adults with TTV MBI progressed with a median time to dementia of 4.82 years (95%CI: 4.48–5.06). Regardless of operational definition, older adults classified as having MBI were more likely to progress to dementia than older adults without MBI (all p<.001). Only 11.1% of those who did not meet the criteria for any MBI progressed to dementia. Sensitivity analysis showed that the prevalence of MBI decreased when using higher MBI cut-off scores [Table S5].

**Figure 3.**
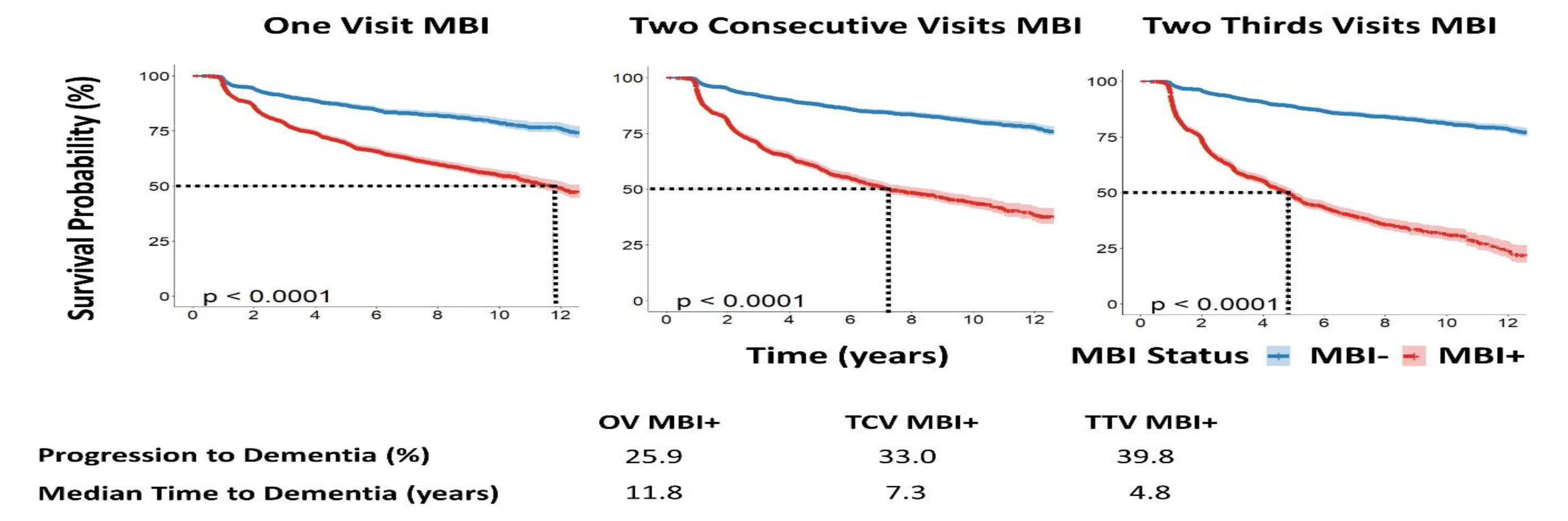
Kaplan-Meier Survival Curves for Three Operational Definitions of MBI. Blue lines indicate the survival curves for older adults without MBI (MBI–) whereas red lines indicate survival curves for older adults with MBI (MBI+) defined using one of three MBI operational definition. Dotted lines in each survival curve indicate the median time to dementia for the MBI+ group in years. All p-values indicated the statistical significance of the relationship between MBI status, using one of the three MBI operational definitions, and incident dementia. Abbreviations: MBI, mild behavioral impairment; OV, One Visit MBI operational definition; TCV, Two Consecutive Visits MBI operational definition; Two-Thirds Visits MBI operational definition. **\***Color should be used for this figure

Univariable Cox proportional hazard regression models showed that dementia risk was lowest in participants with OV MBI, intermediate for TCV MBI, and highest for TTV MBI [Table 2]. Specifically, the HR for OV MBI 2.54 (95%CI: 2.33–2.78, p<.001) compared to participants without MBI, whereas the HRs for TCV and TTV MBI were 4.06 (95%CI: 3.74– 4.40, p<.001) and 6.16 (95%CI: 5.66–6.70, p<.001), respectively. This pattern remained after controlling for age, sex, education, and cognitive status: the adjusted HRs were 1.48 (95%CI: 1.35–1.62, p<.001) for OV MBI, 2.13 (95%CI: 1.96–2.32, p<.001) for TCV MBI, and 2.97 (95%CI: 2.72–3.25, p<.001) for TTV MBI.

**Table 2.** Comparison Between Three MBI Operational Definitions and Their Relationship with Incident Dementia

| **Model** | **HR** | **95% CI** | ***p*** | **C-index** | **AIC** |
| --- | --- | --- | --- | --- | --- |
| Univariable |  |  |  |  |  |
| One Visit MBI | 2.54 | 2.33–2.78 | <.001 | 0.61 | 42412 |
| Two Consecutive Visits MBI | 4.06 | 3.74–4.40 | <.001 | 0.67 | 42140 |
| Two-Thirds Visits MBI | 6.16 | 5.66–6.70 | <.001 | 0.72 | 39709 |
| Multivariable |  |  |  |  |  |
| One Visit MBI | 1.48 | 1.35–1.62 | <.001 | 0.83 | 39355 |
| Two Consecutive Visits MBI | 2.13 | 1.96–2.32 | <.001 | 0.85 | 39357 |
| Two-Thirds Visits MBI | 2.97 | 2.72–3.25 | <.001 | 0.86 | 37357 |

*Note.* All values have been rounded to two decimal places, except for p-values which have been rounded to three decimal places, if appropriate. The outcome variable for all models was incident dementia. The main predictor for all models was MBI status (0 = MBI–, 1 = MBI+) defined using one of three MBI operational definitions. Multivariable models were adjusted for age, sex, education, and cognitive diagnosis (CN/SCD or MCI). C-index and AIC values apply to the corresponding models as a whole. Abbreviations: MBI, mild behavioral impairment, HR, hazard ratio; 95% CI, 95% confidence interval; C-index, concordance index; AIC, Akaike’s information criterion; CN, cognitively normal; SCD, subjective cognitive decline; MCI, mild cognitive impairment.

In both univariable and multivariable Cox regression models, C-index and AIC model metrics indicated that the TTV MBI operational definition was linked to better model performance than TCV and OV operational definitions: C-indices were highest for TTV MBI models, intermediate for TCV MBI models, and lowest for OV MBI models. Correspondingly, AIC was lowest for TTV MBI, intermediate for TCV MBI, and highest for OV MBI models [Table 2]. A sensitivity analysis revealed that these patterns in HR and model performance across MBI operational definitions remained consistent when using higher MBI cut-off scores of ≥2 and ≥3, with marginally larger HRs for higher cut-off scores at the cost of lower similar or slightly worse model performance [Table S5] when compared to an MBI cut-off score of ≥1.

## 4. DISCUSSION

Valuable insight into MBI can be obtained from datasets without MBI-C data by using other NPS assessment tools with the appropriate operational definitions and data cleaning procedures. Here, we outlined and compared three operational definitions for MBI that simply varied in how they addressed the symptom persistence criterion of MBI, while all incorporating the *de novo* symptom emergence criterion by including only those without past psychiatric conditions. The most stringent operational definition of MBI (i.e., TTV) led to lower prevalence estimates of MBI than previously used operational definitions of MBI based on the presence of NPS at one or two consecutive visits. Attendant with TTV-associated group refinement, models produced larger effect sizes and better model metrics for dementia prognostication compared to OV and TCV MBI. These differences suggest that the TTV operational definition yields the greatest specificity for MBI, though comparisons with a gold standard diagnosis of MBI by the MBI-C are warranted.

NPS are common features of dementia, with a five-year period prevalence of 97% in AD patients.^41^ However, when NPS manifest in older adults without dementia and under certain circumstances, they may be an indicator of dementia risk. The MCI criteria may be applied to distinguish chronic cognitive impairment or benign lapses in cognition from cognitive changes suggestive of dementia risk.^42^ Similarly, the MBI criteria play a pivotal role to distinguish chronic and/or recurrent psychiatric conditions and benign alterations in behavior or personality from dementia-related NPS, the latter being sequelae of neurodegenerative disease.^1^ Importantly, MBI represents another tool that can be applied to stratify populations according to dementia risk in clinical and research settings. Clinicians may use MBI to screen patients for dementia risk, with the combination of cognitive and behavioral markers providing greater information on dementia risk than either alone. If appropriate, patients may then be referred to further specialty care, potentially along with more in-depth assessments involving biomarker or neuroimaging tests to confirm a diagnosis. In research studies such as clinical trials, MBI may enrich sample selection by identifying those with a higher likelihood of biomarker positivity, and at greater risk of progression to dementia, thereby reducing screening failures and minimizing variability in outcomes within treatment and control groups.^19^ Altogether, MBI has the potential to be incorporated into patient stratification strategies, which are essential to advance research into dementia, whose causes are multifactorial and whose phenotypes, especially during preclinical or prodromal stages, are heterogeneous.^43,44^ Further research into MBI and its relationship to aspects of dementia is key, therefore, in order to fully utilize MBI in clinical and research settings.

Careful participant exclusion criteria may be applied to address MBI criteria. Participants <50 years of age or with dementia or psychiatric conditions may be excluded during participant selection to ensure that any observed NPS have emerged in later life before a dementia diagnosis, not explained by psychiatric conditions. Satisfying the criterion that requires NPS to persist at least intermittently ≥6 months is more elusive, especially when using NPS assessment tools with one-month reference frames. Our findings highlight the importance of incorporating longitudinal information into MBI case ascertainment, yielding greater effect sizes and better dementia prognostication modeling. The TTV operational definition offers several other advantages over previous MBI operational definitions. By considering a given proportion of visits rather than an absolute number of visits, the TTV definition enables flexible case ascertainment that can become more accurate as more data become available. A dementia-free participant with NPS for both of their only two study visits would be classified as having MBI according to all three operational definitions. However, their classification would change to TTV MBI– if they failed to show NPS for the next three study visits, despite remaining TCV and OV MBI+. Furthermore, the TTV operational definition allows participants to be classified as having MBI even if they do not show NPS for all visits. Participants who fail to report NPS at a study visit due to a variety of potential factors (e.g., a change in informant) may still be classified as MBI+, provided there is sufficient evidence from subsequent study visits that their NPS represents a change from longstanding patterns of behavior (i.e., NPS at more than two-thirds of visits).

This study is not without limitations. First, in the absence of a gold standard clinical diagnosis of MBI in the NACC-UDS, we cannot definitively conclude whether the TTV operational definition enhances specificity for identifying older adults with MBI. Second, given the nature of the NPI and its derivatives that have a one-month reference frame, the TTV operational definition cannot guarantee the persistence of NPS in-between study visits. Third, the TTV operational definition holds little utility in cross-sectional datasets and in clinical practice, where case ascertainment must be performed using information collected at a single visit. Fourth, although the use of all available visit information is a strength of the TTV operational definition, it also means that the amount of information will vary across individuals in studies where participants were followed for various lengths of time. Thus, the MBI-C remains the optimal tool for identifying persons with MBI, as it was developed in accordance with MBI criteria, with a reference frame of six months, allowing MBI status determination at a single visit. The MBI-C is free to use (www.mbitest.org), has been validated in community and clinical samples, and takes approximately seven minutes to administer in-person, by telephone, or online by participants, informants, or clinicians,^5,32,33,45-47^ making it an accessible and scalable tool that can be applied to the growing number of older adults at risk for dementia worldwide.^48^ The TTV operational definition should be used as an alternative to study MBI in datasets where sufficient MBI-C data are not available.

In conclusion, MBI may be implemented in an accessible and scalable manner to help identify older adults at risk of dementia, and can facilitate research and therapies targeting early stages of neurodegenerative disease. While the MBI-C remains the most efficient method to detect and measure MBI in older adults, datasets without MBI-C data may still be used to gain insight into MBI and its relationship with dementia, which is needed to fully utilize MBI in practice. We recommend the TTV operational definition of MBI for datasets that have not implemented the MBI-C. However, for studies based on a comprehensive assessment of risk at a defined time point, the TTV operational definition may not be appropriate or should be modified to be based on the presence of MBI at >2/3 of study visits up to that time point.

## Data Availability

All data produced in the present study are available upon submitting an approved research proposal to the National Alzheimer's Coordinating Center.

## ACKNOWLEDGEMENTS

The NACC database is funded by NIA/NIH Grant U24 AG072122. NACC data are contributed by the NIA-funded ADCs: P50 AG005131 (PI James Brewer, MD, PhD), P50 AG005133 (PI Oscar Lopez, MD), P50 AG005134 (PI Bradley Hyman, MD, PhD), P50 AG005136 (PI Thomas Grabowski, MD), P50 AG005138 (PI Mary Sano, PhD), P50 AG005142 (PI Helena Chui, MD), P50 AG005146 (PI Marilyn Albert, PhD), P50 AG005681 (PI John Morris, MD), P30 AG008017 (PI Jeffrey Kaye, MD), P30 AG008051 (PI Thomas Wisniewski, MD), P50 AG008702 (PI Scott Small, MD), P30 AG010124 (PI John Trojanowski, MD, PhD), P30 AG010129 (PI Charles DeCarli, MD), P30 AG010133 (PI Andrew Saykin, PsyD), P30 AG010161 (PI David Bennett, MD), P30 AG012300 (PI Roger Rosenberg, MD), P30 AG013846 (PI Neil Kowall, MD), P30 AG013854 (PI Robert Vassar, PhD), P50 AG016573 (PI Frank LaFerla, PhD), P50 AG016574 (PI Ronald Petersen, MD, PhD), P30 AG019610 (PI Eric Reiman, MD), P50 AG023501 (PI Bruce Miller, MD), P50 AG025688 (PI Allan Levey, MD, PhD), P30 AG028383 (PI Linda Van Eldik, PhD), P50 AG033514 (PI Sanjay Asthana, MD, FRCP), P30 AG035982 (PI Russell Swerdlow, MD), P50 AG047266 (PI Todd Golde, MD, PhD), P50 AG047270 (PI Stephen Strittmatter, MD, PhD), P50 AG047366 (PI Victor Henderson, MD, MS), P30 AG049638 (PI Suzanne Craft, PhD), P30 AG053760 (PI Henry Paulson, MD, PhD), P30 AG066546 (PI Sudha Seshadri, MD), P20 AG068024 (PI Erik Roberson, MD, PhD), P20 AG068053 (PI Marwan Sabbagh, MD), P20 AG068077 (PI Gary Rosenberg, MD), P20 AG068082 (PI Angela Jefferson, PhD), P30 AG072958 (PI Heather Whitson, MD), P30 AG072959 (PI James Leverenz, MD). NACC funding sources were not involved in the analysis and interpretation of the data, in the writing of the report, or in the decision to submit the article for publication.

## SUPPLEMENTARY GRAPHICS

**Table S1.** One Visit MBI Baseline Participant Characteristics

| Variable | Total | MBI– | MBI+ | *p* |
| --- | --- | --- | --- | --- |
| n | 12771 | 5828 | 6943 |  |
| Age (years) | 73.4 (9.0), 50–104 | 72.4 (9.0), 50–101 | 74.2 (8.9), 50–104 | <.001 |
| Education (years) | 15.7 (3.1), 0–30 | 15.8 (3.0), 0–29 | 15.6 (3.2), 0–30 | .007 |
| Sex (female) | 7313 (57.3) | 3664 (62.9) | 3649 (52.6) | <.001 |
| Diagnosis |  |  |  | <.001 |
| CN | 7967 (62.4) | 4433 (76.1) | 3534 (50.9) |  |
| SCD | 723 (5.7) | 254 (4.4) | 469 (6.8) |  |
| MCI | 4081 (32.0) | 1141 (19.6) | 2940 (42.3) |  |
| MBI Prevalence |  |  |  |  |
| Decreased Motivation | 1365 (10.7) | 0 (0) | 1365 (19.7) | <.001 |
| Affective Dysregulation | 4146 (32.5) | 0 (0) | 4146 (59.7) | <.001 |
| Impulse Dyscontrol | 3934 (30.8) | 0 (0) | 3934 (56.7) | <.001 |
| Social Inappropriateness | 765 (6.0) | 0 (0) | 765 (11.0) | <.001 |
| Psychosis | 293 (2.3) | 0 (0) | 293 (4.2) | <.001 |
| MBI Severity |  |  |  |  |
| Global | 1.3 (1.9), 0–21 | 0 (0), 0–0 | 2.3 (2.1), 1–21 | <.001 |
| Decreased Motivation | 0.1 (0.4), 0–3 | 0 (0), 0–0 | 0.3 (0.6), 0–3 | <.001 |
| Affective Dysregulation | 0.5 (0.9), 0–8 | 0 (0), 0–0 | 0.9 (1.0), 0–8 | <.001 |
| Impulse Dyscontrol | 0.5 (1.0), 0–9 | 0 (0), 0–0 | 0.9 (1.2), 0–9 | <.001 |
| Social Inappropriateness | 0.1 (0.3), 0–3 | 0 (0), 0–0 | 0.1 (0.4), 0–3 | <.001 |
| Psychosis | 0 (0.2), 0–6 | 0 (0), 0–0 | 0.1 (0.3), 0–6 | <.001 |
| Conversion to Dementia | 2437 (19.1) | 648 (11.1) | 1789 (25.8) | <.001 |
| Follow-Up Duration (years) | 3.3 (1.9–6.0) | 3.3 (2.0–6.0) | 3.3 (1.9–6.0) | .53 |

*Note.* Continuous variables are in mean (standard deviation), range, except for follow-up duration which is shown in median (interquartile range). Categorical variables are shown in n (%). P-values indicate the statistical significance of the difference between MBI– and MBI+ groups according to independent samples t-tests or chi-square tests, as appropriate. All values have been rounded to one decimal place, except for p-values which have been rounded to two or three decimal places, if appropriate. Abbreviations: MBI, mild behavioral impairment; CN, cognitively normal; SCD, subjective cognitive decline; MCI, mild cognitive impairment.

**Table S2.**
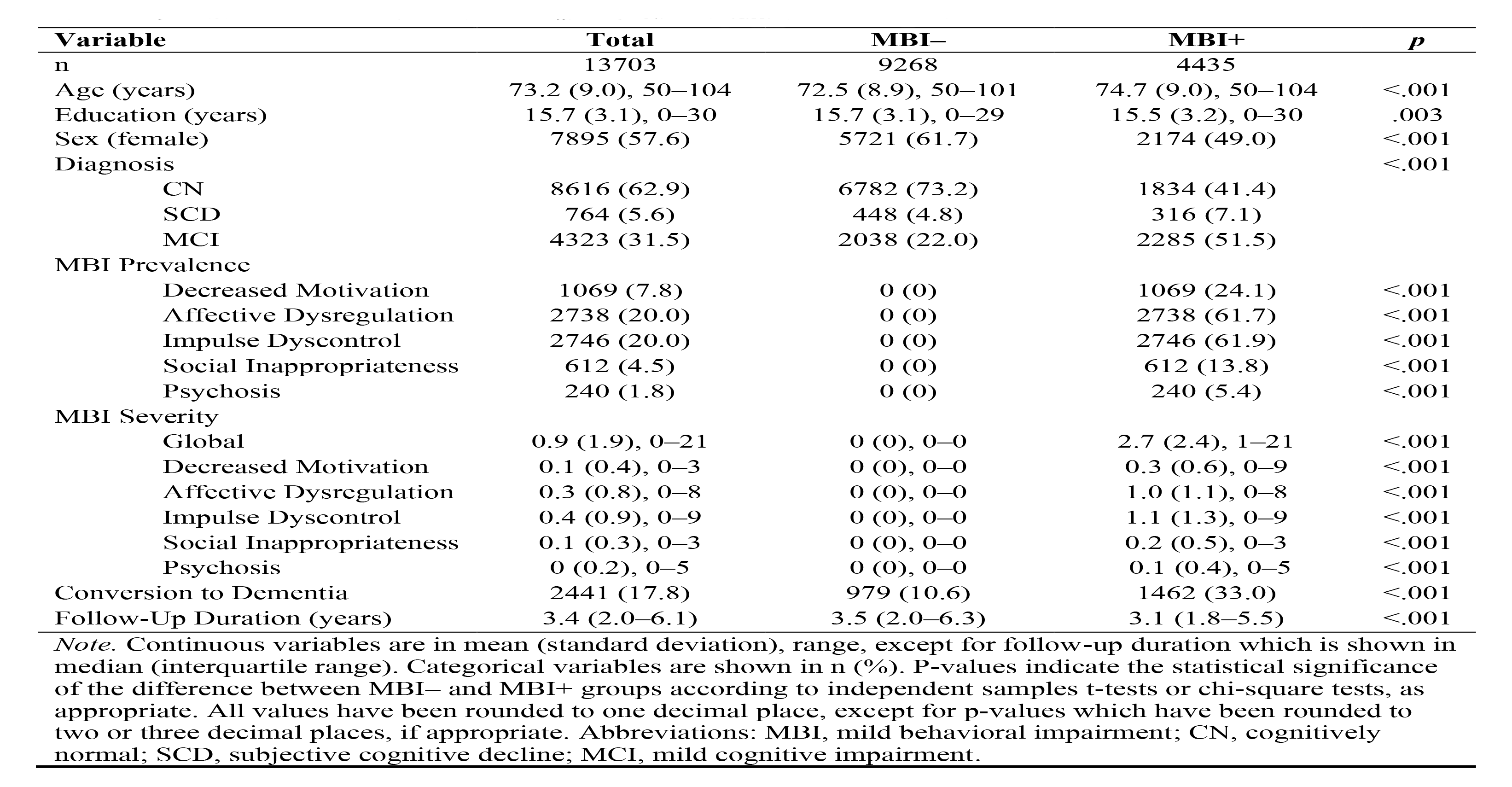
Two Consecutive Visits MBI Baseline Participant Characteristics

| **Variable** | **Total** | **MBI–** | **MBI+** | ***p*** |
| --- | --- | --- | --- | --- |
| n | 13703 | 9268 | 4435 |  |
| Age (years) | 73.2 (9.0), 50–104 | 72.5 (8.9), 50–101 | 74.7 (9.0), 50–104 | <.001 |
| Education (years) | 15.7 (3.1), 0–30 | 15.7 (3.1), 0–29 | 15.5 (3.2), 0–30 | .003 |
| Sex (female) | 7895 (57.6) | 5721 (61.7) | 2174 (49.0) | <.001 |
| Diagnosis |  |  |  | <.001 |
| CN | 8616 (62.9) | 6782 (73.2) | 1834 (41.4) |  |
| SCD | 764 (5.6) | 448 (4.8) | 316 (7.1) |  |
| MCI | 4323 (31.5) | 2038 (22.0) | 2285 (51.5) |  |
| MBI Prevalence |  |  |  |  |
| Decreased Motivation | 1069 (7.8) | 0 (0) | 1069 (24.1) | <.001 |
| Affective Dysregulation | 2738 (20.0) | 0 (0) | 2738 (61.7) | <.001 |
| Impulse Dyscontrol | 2746 (20.0) | 0 (0) | 2746 (61.9) | <.001 |
| Social Inappropriateness | 612 (4.5) | 0 (0) | 612 (13.8) | <.001 |
| Psychosis | 240 (1.8) | 0 (0) | 240 (5.4) | <.001 |
| MBI Severity |  |  |  |  |
| Global | 0.9 (1.9), 0–21 | 0 (0), 0–0 | 2.7 (2.4), 1–21 | <.001 |
| Decreased Motivation | 0.1 (0.4), 0–3 | 0 (0), 0–0 | 0.3 (0.6), 0–9 | <.001 |
| Affective Dysregulation | 0.3 (0.8), 0–8 | 0 (0), 0–0 | 1.0 (1.1), 0–8 | <.001 |
| Impulse Dyscontrol | 0.4 (0.9), 0–9 | 0 (0), 0–0 | 1.1 (1.3), 0–9 | <.001 |
| Social Inappropriateness | 0.1 (0.3), 0–3 | 0 (0), 0–0 | 0.2 (0.5), 0–3 | <.001 |
| Psychosis | 0 (0.2), 0–5 | 0 (0), 0–0 | 0.1 (0.4), 0–5 | <.001 |
| Conversion to Dementia | 2441 (17.8) | 979 (10.6) | 1462 (33.0) | <.001 |
| Follow-Up Duration (years) | 3.4 (2.0–6.1) | 3.5 (2.0–6.3) | 3.1 (1.8–5.5) | <.001 |

*Note.* Continuous variables are in mean (standard deviation), range, except for follow-up duration which is shown in median (interquartile range). Categorical variables are shown in n (%). P-values indicate the statistical significance of the difference between MBI– and MBI+ groups according to independent samples t-tests or chi-square tests, as appropriate. All values have been rounded to one decimal place, except for p-values which have been rounded to two or three decimal places, if appropriate. Abbreviations: MBI, mild behavioral impairment; CN, cognitively normal; SCD, subjective cognitive decline; MCI, mild cognitive impairment.

**Table S3.**
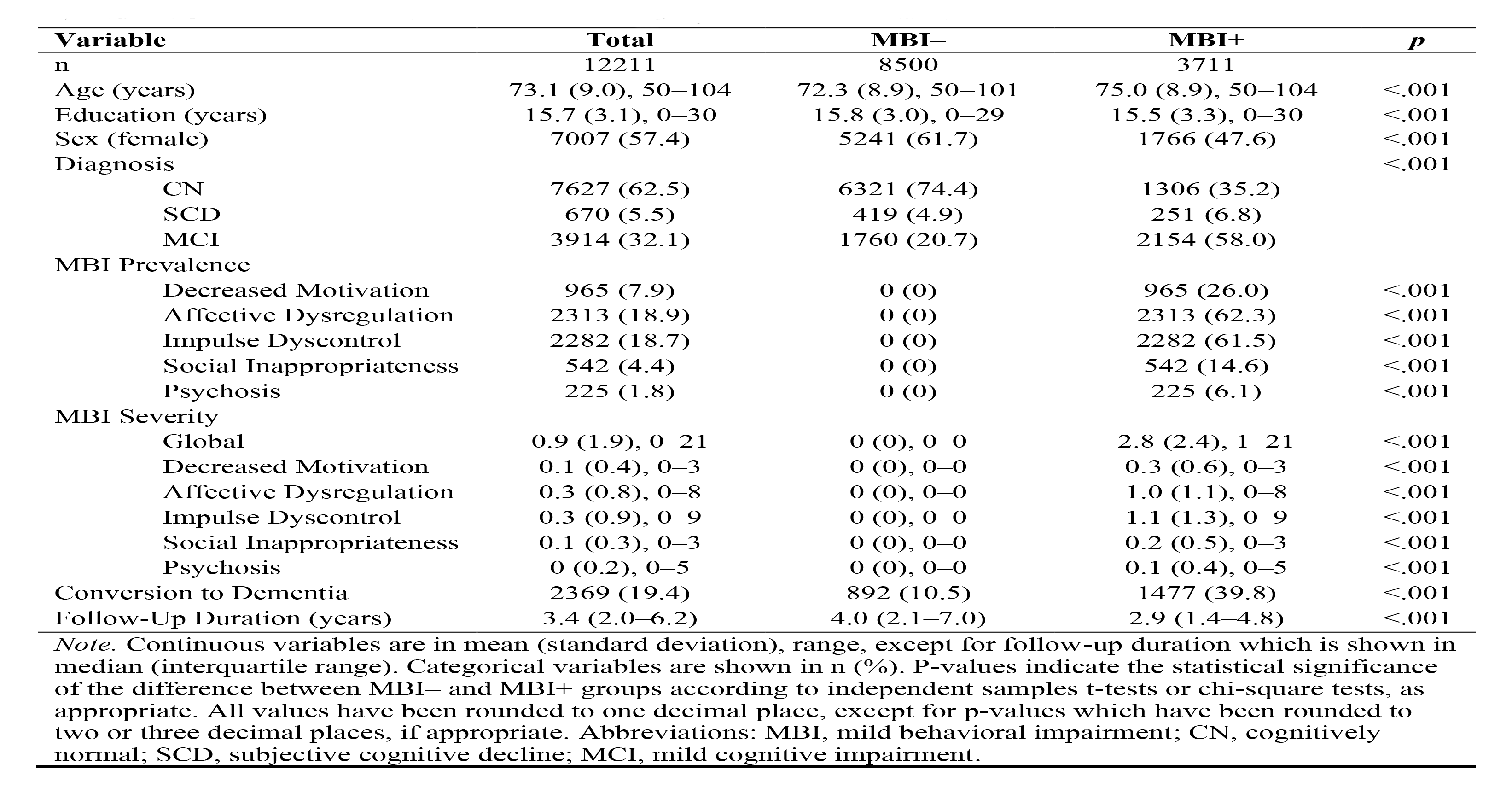
Two-Thirds Visits MBI Baseline Participant Characteristics

| **Variable** | **Total** | **MBI–** | **MBI+** | ***p*** |
| --- | --- | --- | --- | --- |
| n | 12211 | 8500 | 3711 |  |
| Age (years) | 73.1 (9.0), 50–104 | 72.3 (8.9), 50–101 | 75.0 (8.9), 50–104 | <.001 |
| Education (years) | 15.7 (3.1), 0–30 | 15.8 (3.0), 0–29 | 15.5 (3.3), 0–30 | <.001 |
| Sex (female) | 7007 (57.4) | 5241 (61.7) | 1766 (47.6) | <.001 |
| Diagnosis |  |  |  | <.001 |
| CN | 7627 (62.5) | 6321 (74.4) | 1306 (35.2) |  |
| SCD | 670 (5.5) | 419 (4.9) | 251 (6.8) |  |
| MCI | 3914 (32.1) | 1760 (20.7) | 2154 (58.0) |  |
| MBI Prevalence |  |  |  |  |
| Decreased Motivation | 965 (7.9) | 0 (0) | 965 (26.0) | <.001 |
| Affective Dysregulation | 2313 (18.9) | 0 (0) | 2313 (62.3) | <.001 |
| Impulse Dyscontrol | 2282 (18.7) | 0 (0) | 2282 (61.5) | <.001 |
| Social Inappropriateness | 542 (4.4) | 0 (0) | 542 (14.6) | <.001 |
| Psychosis | 225 (1.8) | 0 (0) | 225 (6.1) | <.001 |
| MBI Severity |  |  |  |  |
| Global | 0.9 (1.9), 0–21 | 0 (0), 0–0 | 2.8 (2.4), 1–21 | <.001 |
| Decreased Motivation | 0.1 (0.4), 0–3 | 0 (0), 0–0 | 0.3 (0.6), 0–3 | <.001 |
| Affective Dysregulation | 0.3 (0.8), 0–8 | 0 (0), 0–0 | 1.0 (1.1), 0–8 | <.001 |
| Impulse Dyscontrol | 0.3 (0.9), 0–9 | 0 (0), 0–0 | 1.1 (1.3), 0–9 | <.001 |
| Social Inappropriateness | 0.1 (0.3), 0–3 | 0 (0), 0–0 | 0.2 (0.5), 0–3 | <.001 |
| Psychosis | 0 (0.2), 0–5 | 0 (0), 0–0 | 0.1 (0.4), 0–5 | <.001 |
| Conversion to Dementia | 2369 (19.4) | 892 (10.5) | 1477 (39.8) | <.001 |
| Follow-Up Duration (years) | 3.4 (2.0–6.2) | 4.0 (2.1–7.0) | 2.9 (1.4–4.8) | <.001 |

*Note.* Continuous variables are in mean (standard deviation), range, except for follow-up duration which is shown in median (interquartile range). Categorical variables are shown in n (%). P-values indicate the statistical significance of the difference between MBI– and MBI+ groups according to independent samples t-tests or chi-square tests, as appropriate. All values have been rounded to one decimal place, except for p-values which have been rounded to two or three decimal places, if appropriate. Abbreviations: MBI, mild behavioral impairment; CN, cognitively normal; SCD, subjective cognitive decline; MCI, mild cognitive impairment.

**Table S4.** Demographic Characteristics of Participants With Completed or Missing NPI-Q Data

| Variable | Total | Complete | Missing | *p* |
| --- | --- | --- | --- | --- |
| n | 29231 | 28321 | 910 |  |
| Age (years) | 72.9 (9.3), 50–106 | 72.9 (9.3), 50–104 | 75.4 (10.0), 50–106 | <.001 |
| Education (years) | 15.1 (3.5), 0–30 | 15.1 (3.4), 0–30 | 14.9 (3.9), 0–26 | .37 |
| Sex (female) | 16289 (55.7) | 15766 (55.7) | 523 (57.5) | .28 |
| Diagnosis |  |  |  | <.001 |
| CN | 11929 (40.8) | 11472 (40.5) | 456 (50.1) |  |
| SCD | 1127 (3.9) | 1091 (3.9) | 36 (4.0) |  |
| MCI | 6281 (21.5) | 6128 (21.6) | 153 (16.8) |  |
| Dementia | 9894 (33.8) | 9629 (34.0) | 265 (29.1) |  |

*Note.* Continuous variables are in mean (standard deviation), range, except for follow-up duration which is shown in median (interquartile range). Categorical variables are shown in n (%). P-values indicate the statistical significance of the difference between MBI– and MBI+ groups according to independent samples t-tests or chi-square tests, as appropriate. All values have been rounded to one decimal place, except for p-values which have been rounded to two or three decimal places, if appropriate. Abbreviations: NPI-Q, Neuropsychiatric Inventory Questionnaire; MBI, mild behavioral impairment; CN, cognitively normal; SCD, subjective cognitive decline; MCI, mild cognitive impairment.

**Table S5.** Comparison Between Three MBI Operational Definitions and Their Relationship with Incident Dementia at Variable MBI Cut-off Scores

| Model | HR | 95% CI | *p* | C-index | AIC |
| --- | --- | --- | --- | --- | --- |
| MBI Cut-off $\geq 2$ | | | | | |
| Univariable |  |  |  |  |  |
| OV MBI | 2.71 | 2.50–2.94 | <.001 | 0.62 | 42116 |
| TCV MBI | 4.09 | 3.77–4.45 | <.001 | 0.64 | 42050 |
| TTV MBI | 6.95 | 6.39–7.56 | <.001 | 0.69 | 39922 |
| Multivariable |  |  |  |  |  |
| OV MBI | 1.55 | 1.43–1.69 | <.001 | 0.84 | 39022 |
| TCV MBI | 2.05 | 1.88–2.23 | <.001 | 0.85 | 39145 |
| TTV MBI | 3.08 | 2.83–3.37 | <.001 | 0.87 | 37404 |
| MBI Cut-off $\geq 3$ | | | | | |
| Univariable |  |  |  |  |  |
| OV MBI | 2.83 | 2.61–3.08 | <.001 | 0.61 | 41934 |
| TCV MBI | 4.36 | 3.97–4.79 | <.001 | 0.60 | 41941 |
| TTV MBI | 8.14 | 7.42–8.94 | <.001 | 0.64 | 40368 |
| Multivariable |  |  |  |  |  |
| OV MBI | 1.56 | 1.44–1.70 | <.001 | 0.85 | 38750 |
| TCV MBI | 2.12 | 1.93–2.34 | <.001 | 0.85 | 38859 |
| TTV MBI | 3.38 | 3.07–3.72 | <.001 | 0.87 | 37597 |

*Note.* Using an MBI cut-off of $\geq 2$ , the prevalence was 34.5% ( $n = 4457$ ) for OV MBI, 18.3% ( $n = 2510$ ) for TCV MBI, and 17.3% ( $n = 2169$ ) for TTV MBI. Using an MBI cut-off of $\geq 3$ , the prevalence was 21.6% ( $n = 2826$ ) for OV MBI, 10.4% ( $n = 1425$ ) for TCV MBI, and 9.69% ( $n = 1247$ ) for TTV MBI. All values have been rounded to two decimal places, except for *p*-values which have been rounded to three decimal places, if appropriate. The outcome variable for all models was incident dementia. The main predictor for all models was MBI status (0 = MBI–, 1 = MBI+) defined using one of three MBI operational definitions. Multivariable models were adjusted for age, sex, education, and cognitive diagnosis (CN/SCD or MCI). C-index and AIC values apply to the corresponding models as a whole. Abbreviations: MBI, mild behavioral impairment; HR, hazard ratio; 95% CI, 95% confidence interval; C-index, concordance index; AIC, Akaike’s information criterion; OV, One Visit operational definition; TCV, two consecutive visits operational definition; TTV, two-thirds visits operational definition; CN, cognitively normal; SCD, subjective cognitive decline; MCI, mild cognitive impairment.

